# Biallelic *STAB1* pathogenic variants cause hereditary hyperferritinemia

**DOI:** 10.1101/2022.11.24.22282670

**Authors:** Edoardo Monfrini, Sara Pelucchi, Maija Hollmén, Miro Viitala, Raffaella Mariani, Francesca Bertola, Silvia Majore, Alessio Di Fonzo, Alberto Piperno

**Affiliations:** Dino Ferrari Center, Department of Pathophysiology and Transplantation, University of Milano, Milano, Italy; Foundation IRCCS Cà Granda Ospedale Maggiore Policlinico, Neurology Unit, Milano, Italy; School of Medicine and Surgery, University of Milano-Bicocca, Monza, Italy; MediCity Research Laboratory and InFLAMES flagship, University of Turku, Turku, Finland; Centre for Rare Disease – Disorders of Iron Metabolism, ASST-Monza, San Gerardo Hospital, European Reference Network – EuroBloodNet, Monza, Italy; Cytogenetics and Medical Genetics, ASST-Monza, San Gerardo Hospital, Monza, Italy; Medical Genetics, Department of Molecular Medicine, Sapienza University, San Camillo-Forlanini Hospital, Roma, Italy; Centro Ricerca Tettamanti, Monza, Italy

**Keywords:** Stabilin-1, Ferritin, Hyperferritinemia, Iron, Genetics

## Abstract

Serum ferritin measurement is a routine laboratory test to indirectly evaluate body iron content. However, many additional factors may elevate serum ferritin levels disproportionally to iron stores. Hyperferritinemia is a frequent finding in several conditions, both genetic and acquired. Despite the long history of clinical use, fundamental aspects of the biology of serum ferritin are still unclear. We studied eleven healthy subjects from eight different families presenting unexplained hyperferritinemia without iron overload. To detect the genetic cause of hyperferritinemia we carried out whole-exome sequencing. Immunohistochemistry and flow cytometry assays were performed on patient liver biopsies and monocyte-macrophages to confirm the pathogenic role of the identified candidate variants. Through a combined approach of whole-exome sequencing and homozygosity mapping, we found biallelic candidate *STAB1* variants in ten subjects from seven families. *STAB1* encodes the multifunctional scavenger receptor stabilin-1. Immunohistochemical studies and flow cytometry analyses showed absent or markedly reduced stabilin-1 in patient liver samples, monocytes and monocyte-derived macrophages. We present biallelic *STAB1* mutations as a new cause of inherited hyperferritinemia without iron overload suggesting the existence of new and unexpected function of stabilin-1 in ferritin metabolism.

In conclusion, our findings strongly support biallelic *STAB1* mutations as a novel genetic cause of inherited hyperferritinemia without iron overload and suggest the existence of a new and unexpected function of stabilin-1 in ferritin metabolism.

## Introduction

Hyperferritinemia is a frequent finding in clinical practice and often requires an extensive diagnostic workup. A large spectrum of conditions, both genetic and acquired, associated or not with iron overload, displays high serum ferritin^1-3^. The diagnostic strategy to reveal the cause of hyperferritinemia includes family and personal medical history, biochemical and genetic tests, and evaluation of liver iron by direct (biopsy) or indirect (quantitative magnetic resonance) methods^1^. Despite this complex and time-consuming approach, often the precise etiology remains elusive.

From the molecular point of view, cytosolic ferritin is a 24-subunit protein complex composed of two different subunits, termed H (heavy) and L (light) which are coded by two different genes^4^. Ferritin expression in mammals is regulated by iron through a well-characterized mechanism of coordinated cytosolic post-transcriptional regulation^5^. In addition to iron, ferritin synthesis is regulated by cytokines during development, cellular differentiation, proliferation, and inflammation^6^. Cytokine-dependent control is particularly relevant to inflammation because ferritin is an acute-phase reactant and several pro-inflammatory cytokines stimulate its synthesis^6,7^.

In mammals, a small amount of ferritin is present in a secreted form in serum. It mostly consists of variably glycosylated L-ferritin and trace amounts of H-ferritin^8,9^. Different from cytosolic ferritin, extracellular ferritin is relatively iron-poor^8,9^. For many years, serum ferritin measurement has become a routine laboratory test to indirectly evaluate iron stores, although it is known that many additional factors, including inflammation, infection, and liver diseases, dietary and metabolic abnormalities – all of which may elevate serum ferritin – complicate its interpretation^1,2,10^. Despite this long history of clinical use, fundamental aspects of the biology of serum ferritin are still unclear. For example, tissue of origin, secretory pathway, receptor interactions, clearance, and functions remain topics of active debate^11-13^. It has been hypothesized that ferritin clearance occurs through two different routes: direct uptake by specific ferritin receptors and indirect uptake by ferritin-binding proteins^14^. So far, studies have led to the identification of ferritin receptors such as TIM2, SCARA5, and TFR1^15-17^. TIM2 has no known human ortholog, and specifically bind H-ferritin, SCARA5 is a L-ferritin specific receptor that is required for the development of specific cell types in the embryonic kidney, and TFR1 was also found to bind H-ferritin in human cell lines.

In 2017 we described several Italian subjects with unexplained isolated hyperferritinemia^18^. Four probands had affected siblings, but no affected parents or offspring suggesting the existence of an inherited form of hyperferritinemia without iron overload manifesting as an autosomal recessive trait. We are now able to reveal the presence of biallelic pathogenic variants affecting the multifunctional scavenger receptor stabilin-1 gene (*STAB1*) in most of those patients suggesting that stabilin-1 has a primary role in the regulation of serum ferritin levels in humans.

## Materials and Methods

### Patients

Of the twelve subjects originally reported by Ravasi *et al*.^18^, eleven were available for this study and underwent whole-exome sequencing (WES). They are all Italians and belong to eight different families presenting unexplained hyperferritinemia without iron overload. Their demographic and clinical data have been previously reported^18^ and are summarized in Table 1. These subjects showed levels of serum ferritin markedly higher than expected according to age and gender, ranging from 365 ng/mL to 4654 µg/L. Transferrin saturation (TSAT) was normal (range 20 - 41%), as well as the other laboratory parameters (blood count, liver function tests, metabolic and inflammatory indices). Quantification of liver iron by magnetic resonance showed normal values. Family history collection revealed distant consanguinity in the probands’ parents of family A (#1) and B (#2 and #3). All probands and available relatives gave their written informed consent for genetic testing and research use according to the Institutional Review Board of ASST-Monza, San Gerardo Hospital-Monza (protocol code Gen-CI-001A and GEN-CI-012 approved on 12 October 2018) and for the study “HyFerr” (protocol 2973 approved by the Institutional Ethical Committee on 24 March 2022).

**Table 1.** Iron indices at diagnosis in the 11 patients with hyperferritinemia.

|  | Patient number | Gender | Age (yrs) | Hb (g/dL) | TSAT (%) | s-Ferr (ug/L) | MR-Liver T2* (msec) | MR-LIC (mg/g) |
| --- | --- | --- | --- | --- | --- | --- | --- | --- |
| Family A | #1 | M | 21-25 | 14.9 | 41 | 1313 | 15.2 | 1.67 |
| Family B | #2 | M | 51-55 | 14.5 | 33 | 4654 | 10.6 | 2.42 |
|  | #3 | M | 46-50 | 15.6 | 33 | 2947 | 21.6 | 1.16 |
| Family C | #4 | F | 31-35 | 14.0 | 34 | 987 | 23.8 | 1.05 |
|  | #5 | F | 31-35 | 13.1 | 28 | 365 | 28.6 | 0.87 |
| Family D | #6 | M | 26-30 | 13.1 | 20 | 1970 | 15.2 | 1.67 |
|  | #7 | M | 36-40 | 17.0 | 27 | 2109 | 14.4 | 1.76 |
| Family E | #8 | M | 56-60 | 13.1 | 26 | 3226 | 14.8 | 1.72 |
| Family F | #9 | M | 56-60 | 15.9 | 40 | 1645 | 17.2 | 1.47 |
| Family G | #10 | M | 61-66 | 16.7 | 26.5 | 2903 | 22.0 | 1.14 |
| Family H | #11 | M | 63-66 | 13.6 | 38 | 1154 | 14.8 | 1.72 |

TSAT: Transferrin saturation; s-Ferr: serum ferritin; MR: magnetic resonance; LIC: liver iron concentration (normal values: men $\leq 1.8$ , in women $\leq 1.2$ mg/g).

### Genetic methods

WES was carried on using the Nextera Rapid Capture Exome Library kit (Illumina) on the Illumina NextSeq 550 System (San Diego, California, USA). Reads alignment and variant calling/annotation were performed using BWA-MEM algorithm and Picard-GATK4 tools (Broad Institute). Homozygosity mapping was performed starting from WES data using the AutoMap online tool (https://automap.iob.ch/) (Parameters: DP=8, percaltlow=.25, percalthigh=.75, binomial=.000001, maxgap=10, window=7, windowthres=5, minsize=2, minvar=25, minperc=88, chrX=No, extend=1)^19^. WES variants were prioritized using the following criteria: allele frequency (MAF) ≤ 0.001, quality score (QS) ≥ 30, protein impact, *in silico* prediction of pathogenicity (CADD, DANN, Polyphen2, SIFT-PROVEAN, MutationTaster)^20-24^, homozygous or compound heterozygous status, and genotype identity in affected siblings (when applicable). The candidate variants were validated by Sanger sequencing in the probands and available parents.

### Immunohistochemistry

Three patients (#9, #2 and #3) underwent liver biopsy for diagnostic purposes in the past. Blank samples were obtained from Formalin-Fixed Paraffin-Embedded (FFPE) specimens. The control liver samples were derived from subjects with hypertransaminasemia whose histology was normal (kindly provided by the hepatology outpatient clinic of the ASST-Monza, San Gerardo Hospital). FFPE 4-5 µm slices were stained for stabilin-1 using Ventana Benchmark Ultra (Roche Diagnostics, Basel, Switzerland) with an anti-stabilin-1 primary antibody (clone 4G9, sc-293254, Santa Cruz, Dallas, TX, USA) at 1:100 dilution combined with UltraView Universal DAB Detection Kit (Roche Diagnostics, Basel, Switzerland). Anti-Vascular adhesion protein 1 (VAP1) antibodies were used to check for liver integrity of the analyzed samples (polyclonal rabbit anti-serum against human VAP1, produced in-house and used 1:5000).

### Flow cytometry

Mononuclear cells were isolated by Ficoll gradient centrifugation (GE Healthcare, Amersham Biosciences Europe GmbH, Freiburg, Germany) from 40 ml of blood of patients (#2, #3, #1, and #10) and healthy controls, and human monocytes were isolated from peripheral blood mononuclear cells by magnetic enrichment using CD14+ microbeads according to the manufacturer’s instructions (Miltenyi Bergisch Gladbach, Germany). Patient monocytes were used as such or differentiated (patients #2 and #1) into macrophages and M2 polarized as previously described^25^. For flow cytometry, cells were stained with anti-stabilin-1 (clone 9-11, InVivo Biotech Hennigsdorf, Germany) conjugated with Alexa Fluor 647 in-house, anti-CD14-Pacific Blue (558121, BD, Vernon Hills, Illinois, USA) or isotype-matched irrelevant antibody. 7-AAD was used as a viability dye (Invitrogen, 00-6993-50). Samples were acquired on the LSRFortessa (BD, Vernon Hills, Illinois, USA) and analyzed with FlowJo v. 10.8.1 (BD, Vernon Hills, Illinois, USA).

### Data Sharing Statement

For original data, please contact for clinical and genetic data, for whole exome studies, and for functional studies.

## Results

### Genetic study

In the hypothesis of a recessive phenotype, we searched for rare homozygous or compound heterozygous variants in WES data. Based on patients’ phenotype we did not expect and did not find pathogenic variants in the hemochromatosis genes or genes associated with hereditary iron overload. We also confirmed the negative findings obtained with Sanger sequencing in L-ferritin and ferroportin genes^18^. Patients of families A (#1) and B (#2 and #3) showed a higher homozygosity rate than controls in line with the known distant consanguinity of their parents. Homozygosity mapping showed a single chromosomal region of homozygosity overlap in the three patients on chromosome 3 (hg19, Chr3: 50,095,103-54,420,721 bp) (Figure 1). Looking for rare non-synonymous variants in this locus we found two homozygous variants in the *STAB1* gene: a missense variant (p.Ser2244Pro) in subject #1 and a deletion/insertion variation (c.5612_5646delinsG) leading to a premature stop codon (p.Gly1871*) in subjects #2 and #3. Interestingly, *STAB1* gene display mild-to-moderate intolerance to missense and loss of function variants (o/e = 0.92 and 0.68, respectively -gnomAD). In addition, a recent meta-analysis of genome-wide association studies (GWAS) found deleterious *STAB1* variants associated with ferritin levels^26^. Therefore, the *STAB1* gene was prioritized as the candidate etiological gene and was analyzed in the remaining probands with unexplained hyperferritinemia. Remarkably, we found *STAB1* variants in seven of the remaining eight subjects with hyperferritinemia. In details, six carried rare compound heterozygous variants in *STAB1*: p.Glu348Lys and p.Trp2443* in subjects #4 and #5 (Family C), p.Gly723_Phe725del and p.Arg1122Profs*37 in subjects #6 and #7 (Family D), p.Ser2244Pro and p.Tyr2339Cys in subject #8 (Family E), p.Cys120Ser and the rare synonymous variant p.Cys784= in subject #9. Last, subject #10 resulted homozygous for the same p.Ser2244Pro variant found in probands #1 and #8. Interestingly, the three patients carrying this variant (#1, #8, and #10) originated from a restricted geographical area in the south of the Apulia region suggesting a founder effect. All the variants were confirmed by Sanger sequencing. The available parents, who presented normal serum ferritin, were heterozygous carriers of the *STAB1* variants, demonstrating biallelic status in the probands. Table 2 summarizes the identified *STAB1* variants and Supplementary figures 1 and 2 show the family pedigrees and *STAB1* variants visualization in the .bam files of the affected subjects, respectively.

**Figure 1.**
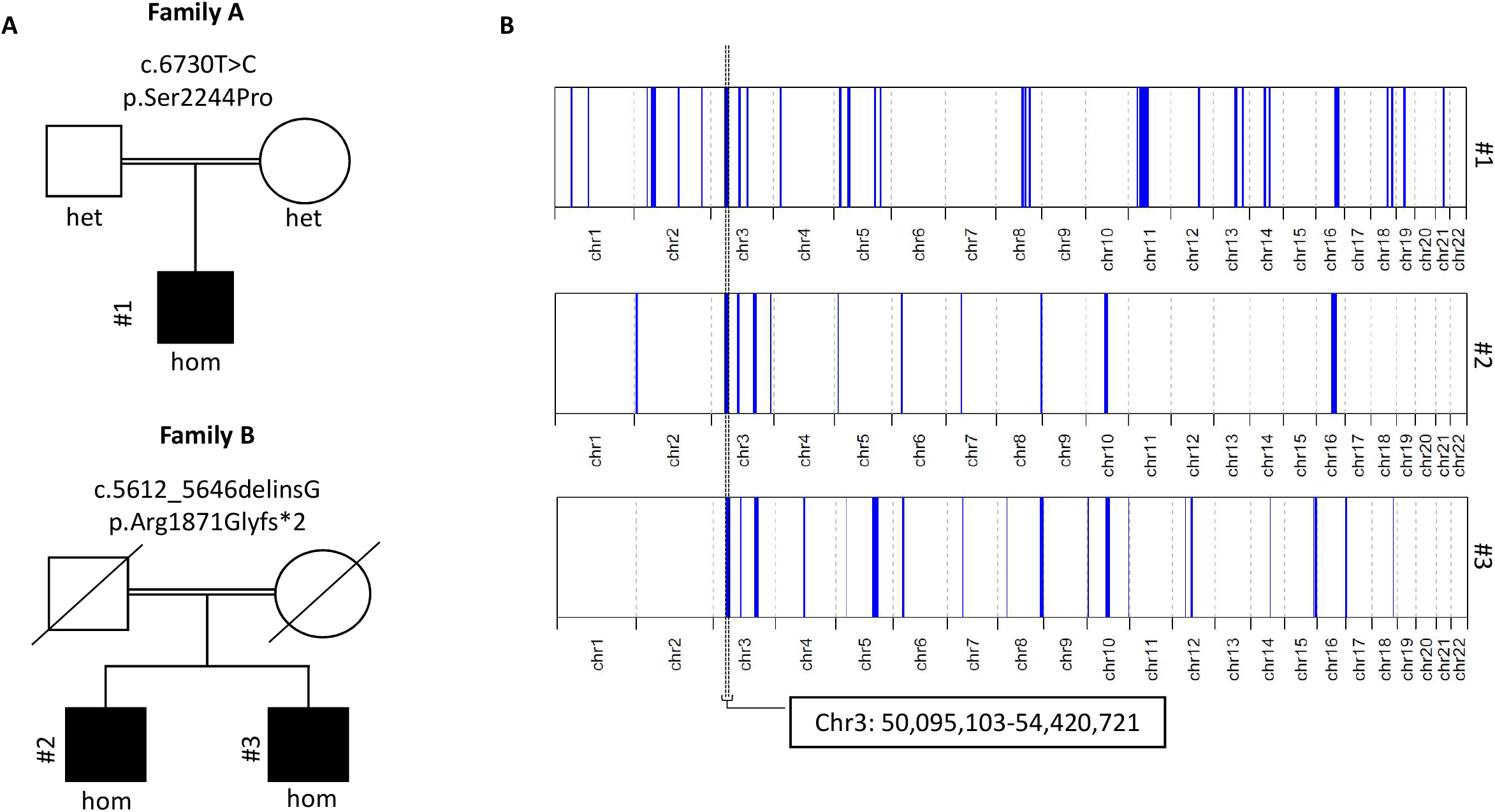
(A) Pedigrees of the two families with history of distant consanguinity carrying the candidate *STAB1* variants. Black symbols denote affected status (hom = homozygous, het = heterozygous). (B) Homozygosity mapping analysis in patients #1 (Family A), #2 and #3 (Family B), performed with AutoMap online tool (https://automap.iob.ch/)^19^, revealed a single chromosomal region of homozygosity overlap on chromosome 3 (hg19, Chr3:50,095,103-54,420,721 bp), where *STAB1* gene is localized (chr3:52,529,354-52,558,511).

**Table 2.** Genetic characterization of the identified *STAB1* variants

| **Patients** | **Variants (cDNA)** | **Variants (AA)** | **Allelic status** | **In silico predictions** | **Mammalian conservation** | **rs number (dbSNP)** | **MAF (gnomAD_exomes)** |
| --- | --- | --- | --- | --- | --- | --- | --- |
| #1 | c.6730T>C | p.Ser2244Pro | Homozygous | Deleterious (5/5) | Yes | rs141939118 | 0.000028 |
| #2 and #3 | c.5612_5646delinsG | p.Gly1871* | Homozygous | Deleterious (2/2) | NA | NA | NA |
| #4 and #5 | c.1042G>A | p.Glu348Lys | Heterozygous | Deleterious (4/5) | Yes | rs754318051 | 0.000005 |
|  | c.7328G>A | p.Trp2443* | Heterozygous | Deleterious (3/3) | NA | rs748728975 | 0.000004 |
| #6 and #7 | c.2167_2175del | p.Gly723_Phe725del | Heterozygous | Likely deleterious (1/2) | Yes | NA | NA |
|  | c.3364dup | p.Arg1122Profs*37 | Heterozygous | Deleterious (2/2) | NA | rs563085224 | 0.000064 |
| #8 | c.6730T>C | p.Ser2244Pro | Heterozygous | Deleterious (5/5) | Yes | rs141939118 | 0.000028 |
|  | c.7016A>G | p.Tyr2339Cys | Heterozygous | Deleterious (5/5) | Yes | rs1484029620 | 0.000014 |
| #9 | c.358T>A | p.Cys120Ser | Heterozygous | Deleterious (5/5) | Yes | rs778572255 | 0.000004 |
|  | c.2352C>T | p.Cys784= | Heterozygous | Not deleterious (0/2) | No | rs898828640 | 0.000012 |
| #10 | c.6730T>C | p.Ser2244Pro | Homozygous | Deleterious (5/5) | Yes | rs141939118 | 0.000028 |

AA: amino acid; MAF: minor allele frequency; NA: Not Applicable. The following tools have been used for the in silico predictions: CADD, DANN, Polyphen2, SIFT-PROVEAN, MutationTaster, SpliceAI, VarSEAK, Human Splicing Finder (when applicable).

### Immunohistochemistry and flow cytometry

To understand how the genetic variants affected stabilin-1 protein levels, we stained liver biopsy samples from patients and controls with an anti-stabilin-1 antibody. In the liver, stabilin-1 is known to be expressed by liver sinusoids but not by Kupffer cells^27^. Patients #9, #2 and #3 showed no immunoreactivity with anti-stabilin-1 compared to control liver where high signal was detected in the liver sinusoids (Figure 2). All the samples stained normally with anti-vascular adhesion protein 1 (VAP-1) antibody used as control staining confirming the integrity of the samples (Supplementary Figure 3). Thus, the staining results suggest that the identified stabilin-1 genetic variants lead to complete loss of the expressed protein in the liver.

**Figure 2.**
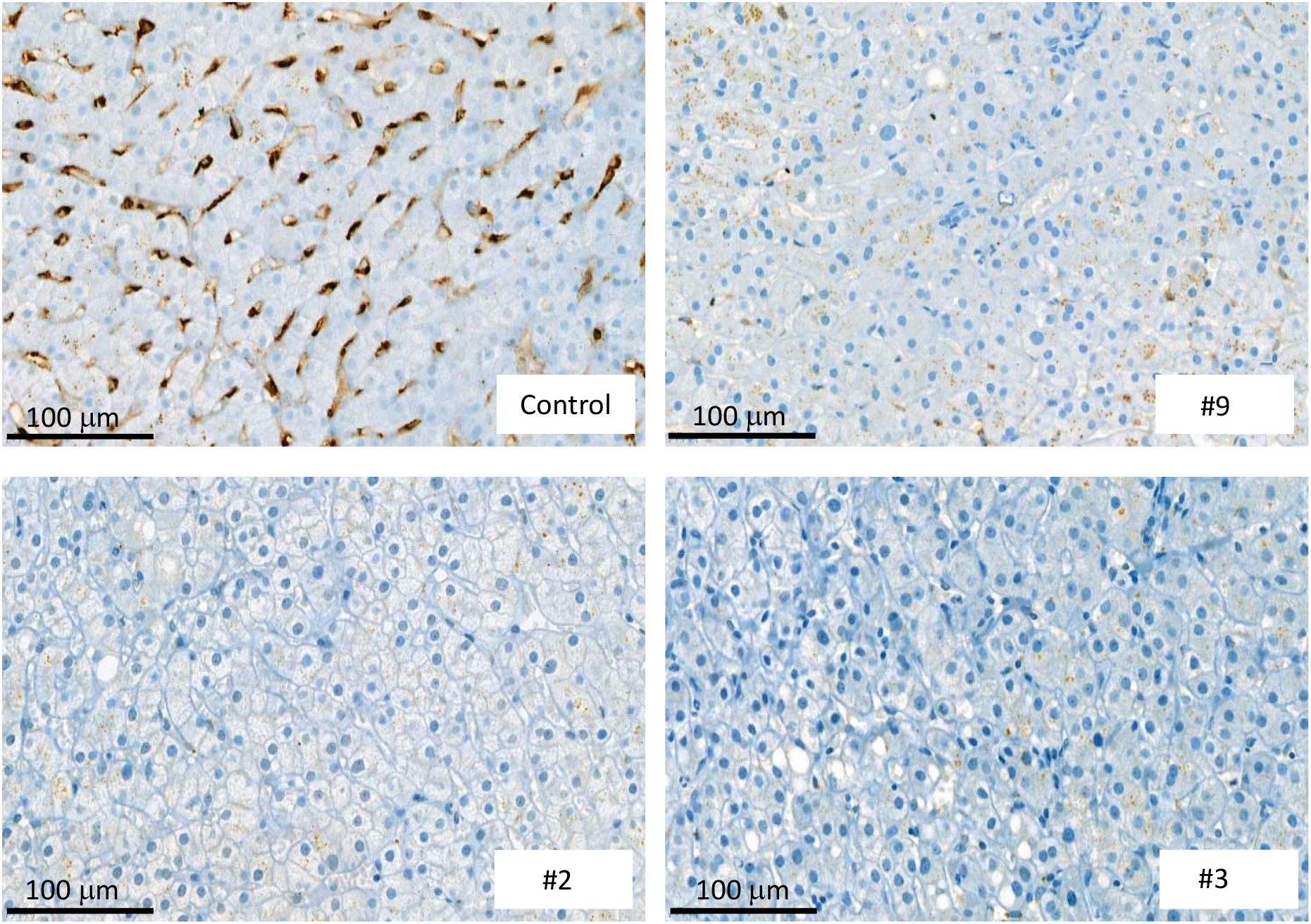
Anti-Stabilin-1 staining of liver biopsy samples from a healthy control and patients #9, #2, and #1 (40X).

To validate this finding on a more accessible cell population, we next investigated stabilin-1 expression levels on peripheral monocytes (#2, #3, #1, and #10) and monocyte-derived macrophages obtained from patients #2 and #1. Consistently, we observed very little expression of stabilin-1 on CD14+ monocytes and macrophages (Figure 3).

**Figure 3.**
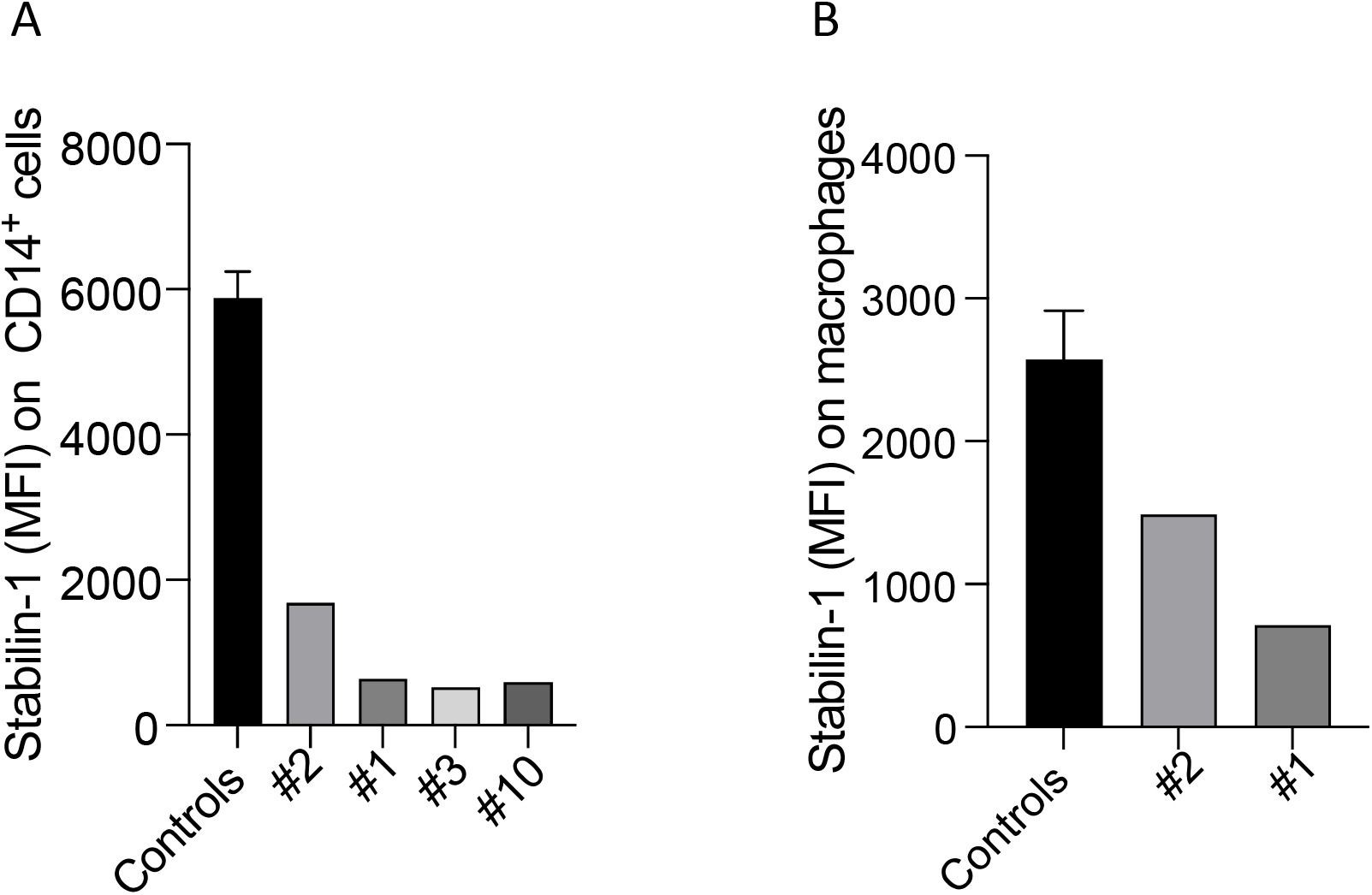
Flow cytometric analysis of cell-surface stabilin-1 on CD14^+^ monocytes in controls (n=3) and in patients #2, #1, #3 and #10 (A), and on monocyte-derived macrophages from controls (*n*=3) and patients #2 and #1 (MFI: median fluorescence intensity) (B).

## Discussion

There are several lines of evidence supporting the pathogenic role of the identified *STAB1* variants and their relations with hyperferritinemia. First, the co-segregation of biallelic *STAB1* variants with hyperferritinemia in three unrelated families (#2, #3, #4, #5, #7and #6) is a major proof of pathogenicity. Second, the nonsense variants (p.Gly1871*, p.Trp2443*, p.Arg1122Profs*37) are highly deleterious as they are expected to cause a premature stop codon leading to a truncated stabilin-1 protein. Third, the identified single nucleotide variants and small indels are extremely rare (only identified in the heterozygous status in the population database gnomAD)^28^ affecting amino acids highly conserved across mammalian species and predicted to be deleterious by the majority of *in silico* tools, except for the p.Cys784= variant carried by patient #9 (Table 2). This variant deserves separate consideration since it is synonymous, it does not affect a conserved nucleotide, and *in silico* prediction tools predict it to be benign. A splicing disruption effect can be hypothesized; however, *in silico* prediction tools do not consistently support this hypothesis (i.e., SpliceAI, VarSEAK, Human Splicing Finder). Unfortunately, RNA from subject #9 was not available for *STAB1* transcript analyses. Alternatively, another unidentified non-coding pathogenic *STAB1* variant may explain the depletion of stabilin-1 in this subject. However, liver immunohistochemistry stabilin-1 measurement (Figures 2) indicate that #9 is unable to express the protein as observed for patients #2 and #3 carrying nonsense mutations, indicating a deleterious *STAB1* genotype in this subject. Fourth, the analysis of in-house exomes of 500 subjects without hyperferritinemia (Italian neurological patients) did not reveal any carrier of these specific variants or rare biallelic *STAB1* variations (personal data). Fifth, a recent GWAS meta-analysis found an association between four uncorrelated rare *STAB1* variants (p.Glu117*, p.Gly189Ser, p.Glu527Lys, and p.Ser1089Gly) and increased serum ferritin, and in line with our observations, those *STAB1* variants did not associate with increased iron levels and transferrin saturation^26^. Sixth, immunohistochemistry studies displayed a striking reduction of stabilin-1 protein in hepatic sinusoidal cells of *STAB1* mutation carriers compared to controls. Seventh, patient-derived blood monocytes and macrophages displayed a marked decrease of stabilin-1 on cell surfaces.

Stabilin (or FEEL/CLEVER/HARE) receptors belong to class H scavenger receptors that consists of two members, Stabilin-1 and Stabilin-2^29,30^. The stabilins are enigmatic proteins whose physiological functions are still not entirely understood^31^. They comprise a large extracellular N-terminus of multiple epidermal growth factor (EGF)/EGF-like domains, seven fasciclin-1 domains, an X-link domain, and a short intracellular C-terminal domain, linked by a transmembrane region^30^. Their extracellular domains share 55% similar homology, but their short intracellular domains are highly diverse, which results in differential abundance in different tissues and cells^31,32^. More specifically, stabilin-1 is primarily expressed on sinusoidal endothelial cells of the liver, spleen, adrenal cortex and lymph nodes, and tissue macrophages^30,32,33^. The expression profile of this transmembrane protein in professional scavenging cells (sinusoidal endothelial cells and macrophages) strongly supports its function as a scavenger receptor. However, unexpectedly for scavenger receptors, stabilin-1 is the first example of a macrophage-specific receptor that mediates endocytic clearance, intracellular sorting and transcytosis^34^. Due to the multifaceted function of stabilin-1, different hypotheses can be drawn to explain our findings. First, stabilin-1 may have a role as a scavenger receptor for serum ferritin at the surface of macrophages and/or liver sinusoidal endothelial cells. There is general agreement that cells uptake ferritin through two routes: direct uptake by ferritin receptors and indirect uptake by ferritin-binding proteins^14^. It was shown that SCARA5, which belongs to scavenger receptor class A, mediates the uptake of ferritin iron in specific cell types in the developing kidney in mice^16^, and that SCARA5-transfected cells can bind or internalize human L-ferritin and H-ferritin ^35^. Stabilin-1, which is able to endocytose ligands such as low-density lipoproteins, Gram-positive and Gram-negative bacteria, and advanced glycosylation end products^30,36^, might be another scavenger receptor for ferritin in other cell types. Second, stabilin-1 might control ferritin metabolism through more complex mechanisms that concern its role in the regulation of the macrophage secretome^34^. Stabilin-1 shuttles between endosomal compartment and biosynthetic compartment and transports newly synthesized stabilin-interacting proteins to the lysosomal secretory pathway. Interestingly, it has been previously proposed that serum ferritin is secreted through the nonclassical lysosomal secretory pathway in mice, specifically through secretory lysosomes^11^. This is supported by the recent findings showing that specific defects of proteins involved in early or later stages of endo-lysosomal trafficking have opposite effects on ferritin secretion^37^. Both hypotheses are in agreement with what we have previously observed in these patients who showed elevated serum ferritin levels in the face of normal ferritin concentrations in lympho-monocytes suggesting altered ferritin secretion or clearance ^18^.

In conclusion, we present biallelic *STAB1* mutations as a novel genetic cause of hereditary hyperferritinemia without iron overload. Although such relationship is strongly supported by the data, the underlying mechanisms linking stabilin-1 to ferritin remain to be defined and requires further studies to be clarified. Our findings also indirectly suggest that this function mainly attains to stabilin-1 in humans, since stabilin-2 protein appears to be unable to compensate for the defective stabilin-1. At present, there is no evidence that *STAB1* mutations can lead to clinical manifestations other than hyperferritinemia as all the patients reported here are healthy, although later manifestations cannot be excluded considering the role of stabilin-1 in inflammatory response^38^. Last, our results support *STAB1* sequencing as a new test in the differential diagnostic set up of hyperferritinemia allowing more precise diagnosis and avoiding useless and expensive investigations in the clinical practice.

## Abbreviations

ELISA: Enzyme-Linked ImmunoSorbent Assay
FFPE: Formalin-Fixed Paraffin-Embedded
GWAS: Genome-Wide Association Studies
STAB1: STABilin-1 gene
TSAT: Transferrin SATuration
WES: Whole-Exome Sequencing

## Acknowledgement

We thank Dr. Antonio Ciaccio (Hepatology Unit ASST-Monza, San Gerardo Hospital) for providing samples of control liver biopsies and Dr. Nicola Zucchini and the technicians of the Pathology Unit ASST-Monza, San Gerardo Hospital for providing slices of PPFE liver samples of patients and controls. We thank patients and parents involved in the study, and the Associazione per lo Studio dell’Emocromatosi e delle Malattie da Sovraccarico di Ferro (Association for the Study of Hemochromatosis and Iron Overload Disorders) + Fe – ONLUS, Monza for their helpfulness.

## Authorship and conflict-of-interest statements

Contribution: S.P. and A.P. designed research; E.M., S.P. M.H., M.V., F.B., and A.D.F. performed experiments; E.M., S.P., R.M., S.M., A.D.F., and A.P. collected and/or analyzed data; E.M., S.P., M.H., S.M., A.D.F., and A.P. wrote the manuscript; and all authors revised the manuscript.

