## Supplementary Figures for "Biallelic *STAB1* pathogenic variants cause hereditary hyperferritinemia"

**Supplementary Figure 1.** Family pedigrees of the patients with hyperferritinemia carrying *STAB1* variants.

**Supplementary Figure 2.** Visualization of the identified *STAB1* variants on WES files (bam format) using Integrative Genomics Viewer (IGV).

**Supplementary Figure 3.** Example staining of VAP-1 in the liver of patient PATIENT#3 (10x and 40x).

**Supplementary figure 1**

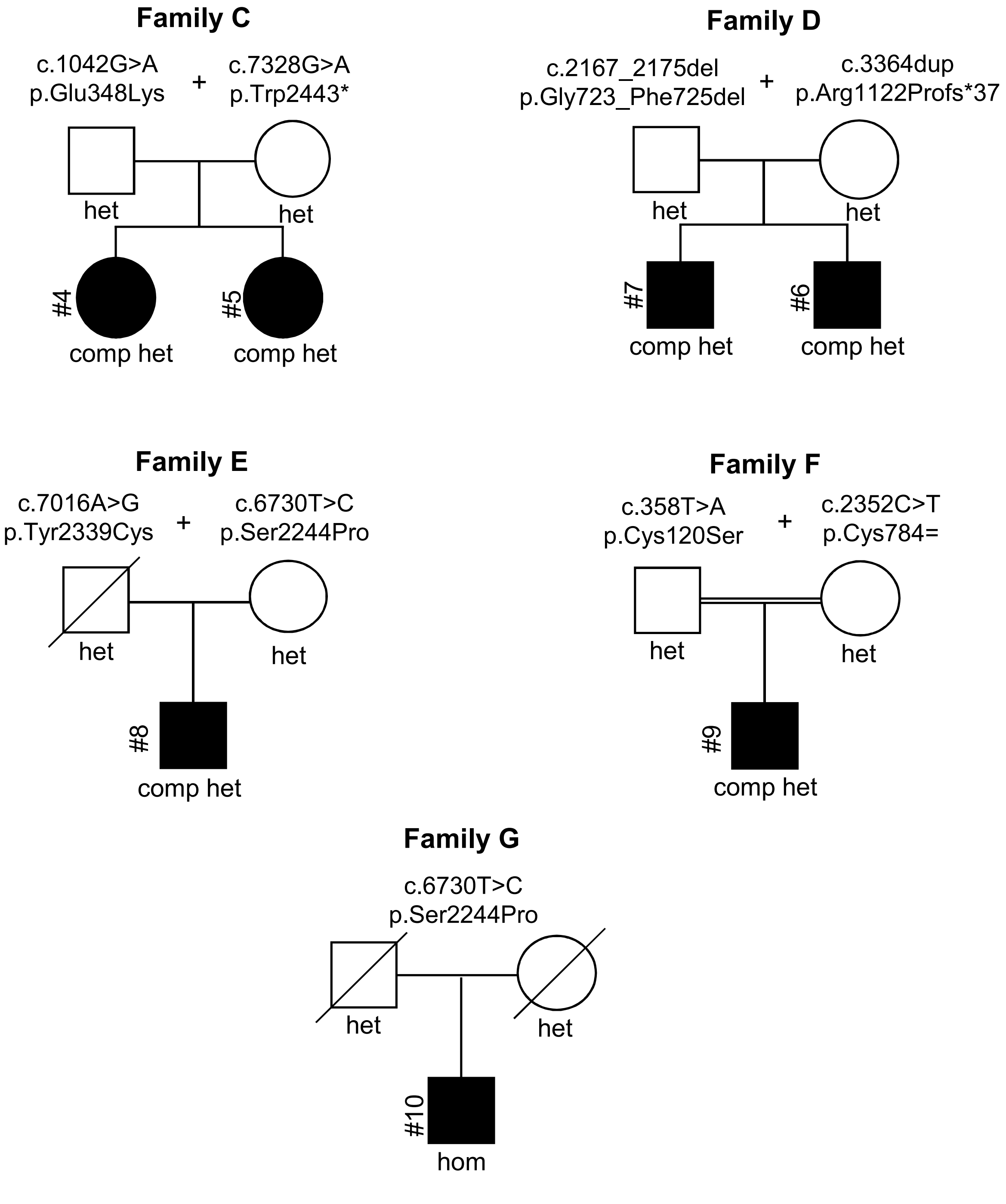

**Supplementary figure 2**

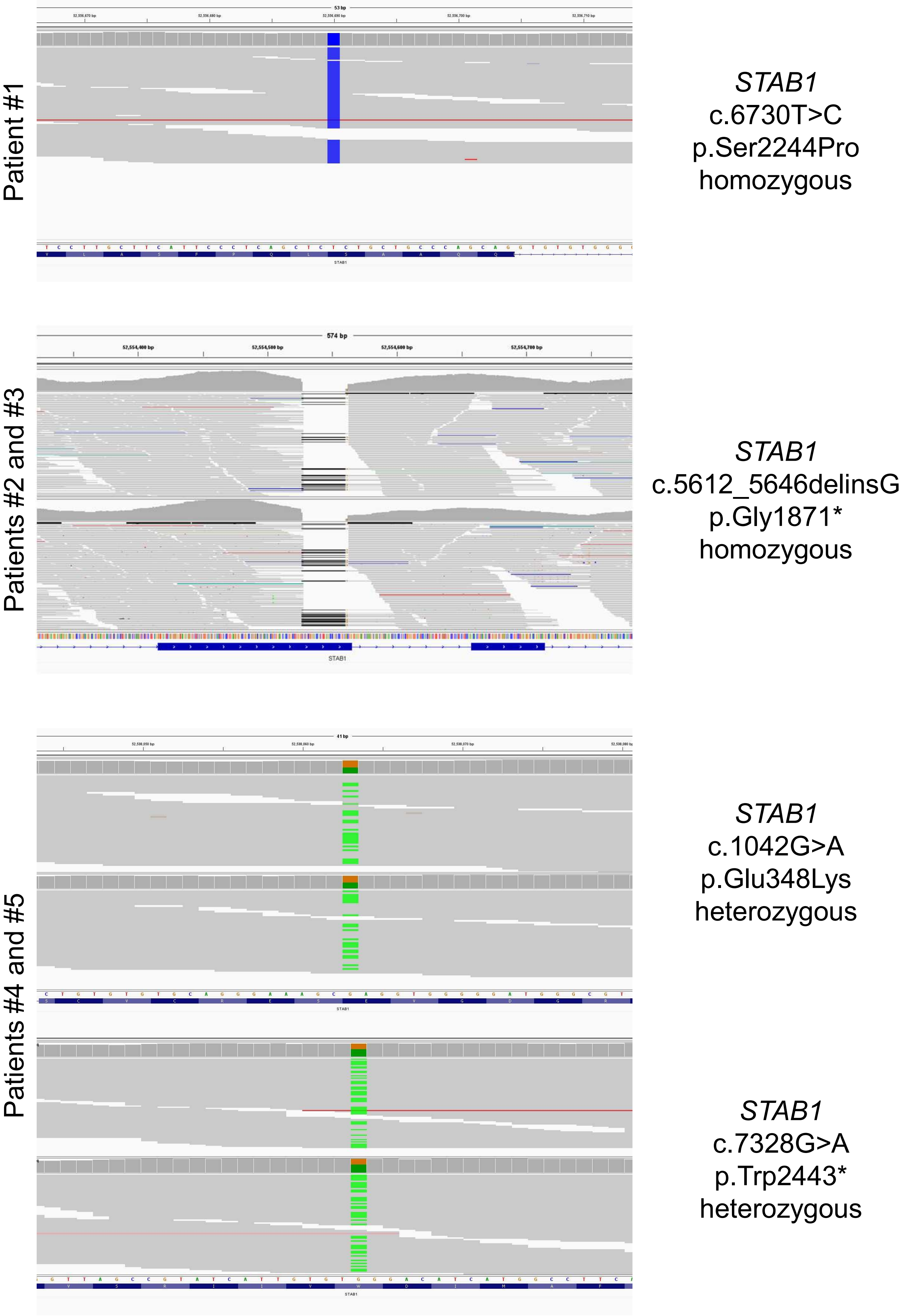

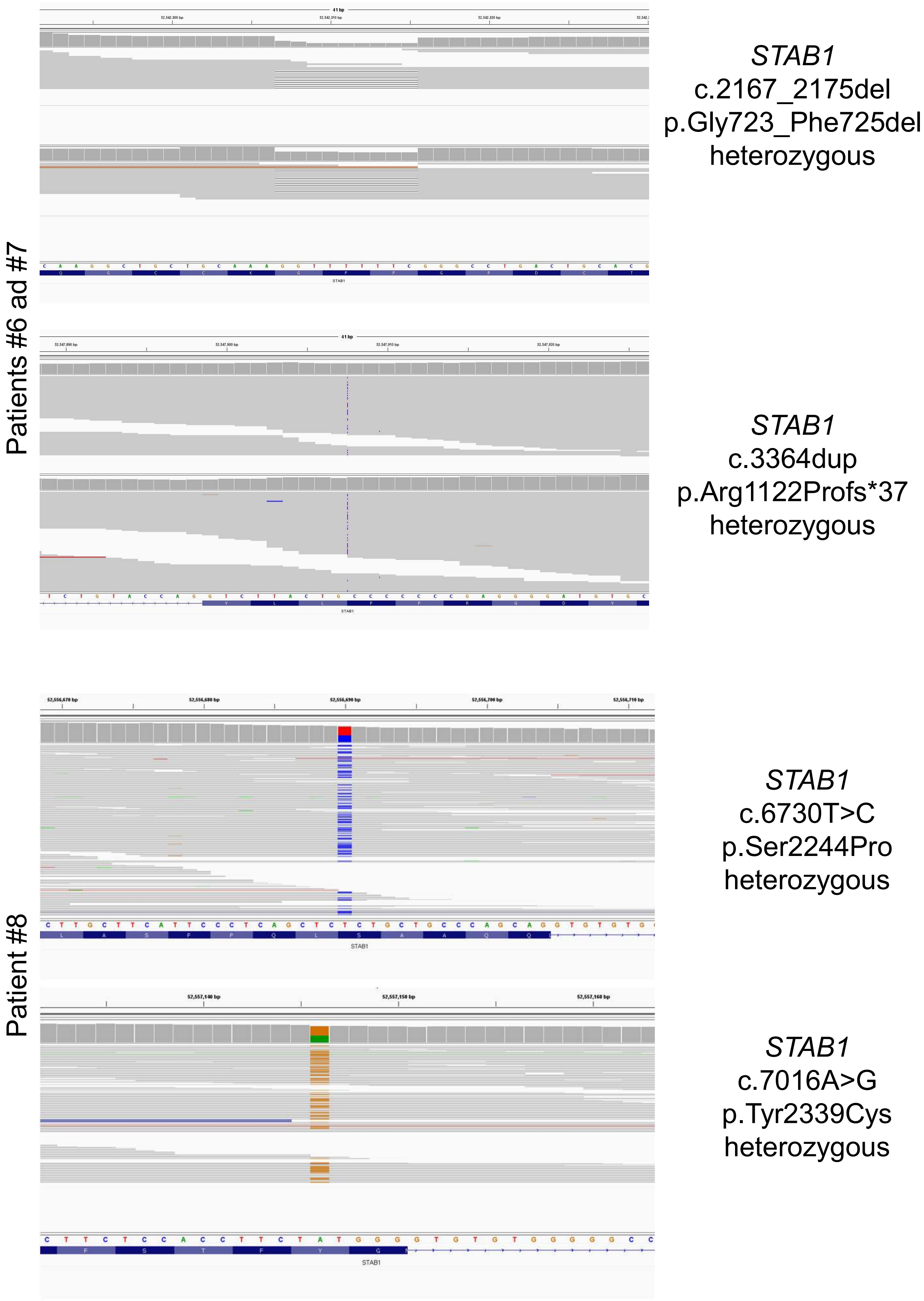
Patients #6 ad #7
STAB1
c.2167_2175del
p.Gly723_Phe725del
heterozygous
STAB1
c.3364dup
p.Arg1122Profs*37
heterozygous
Patient #8
STAB1
c.6730T>C
p.Ser2244Pro
heterozygous
STAB1
c.7016A>G
p.Tyr2339Cys
heterozygous

Patient #9

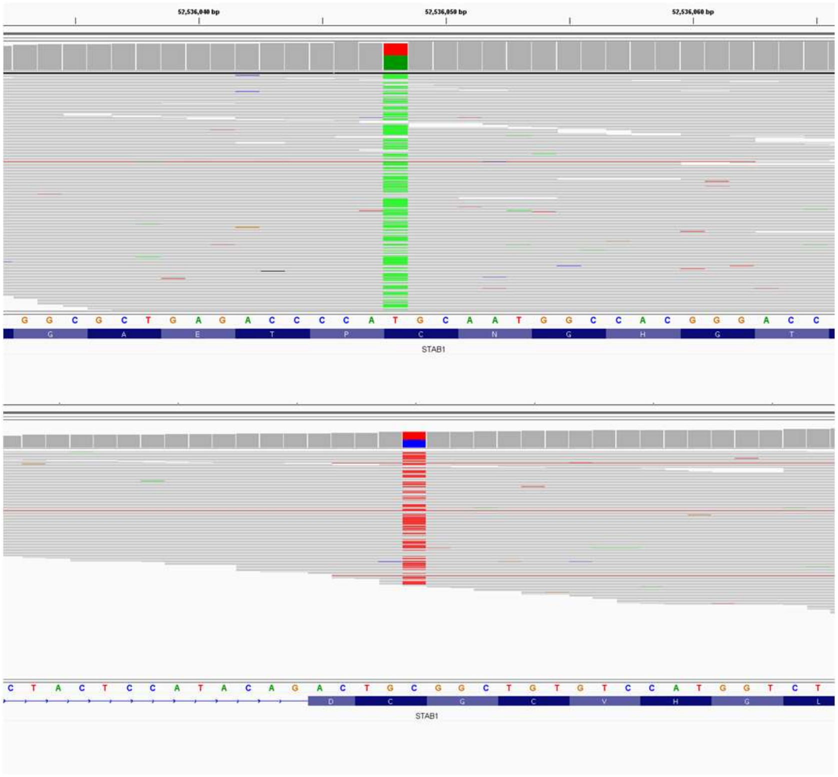

*STAB1*
c.358T>A
p.Cys120Ser
heterozygous

*STAB1*
c.2352C>T
p.Cys784=
heterozygous

Patient #10

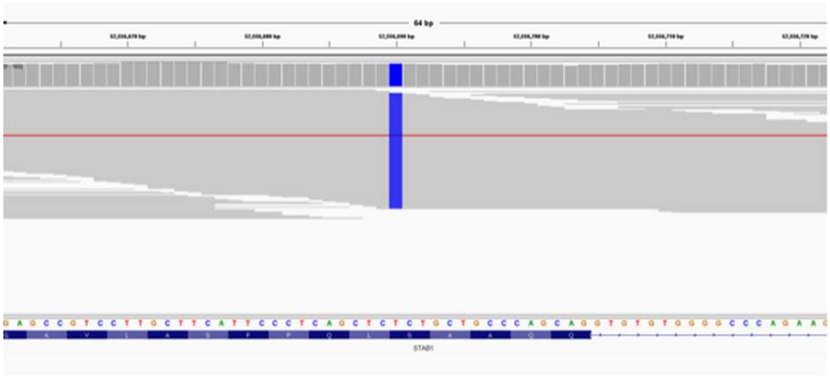

*STAB1*
c.6730T>C
p.Ser2244Pro
homozygous

**Supplementary figure 3**

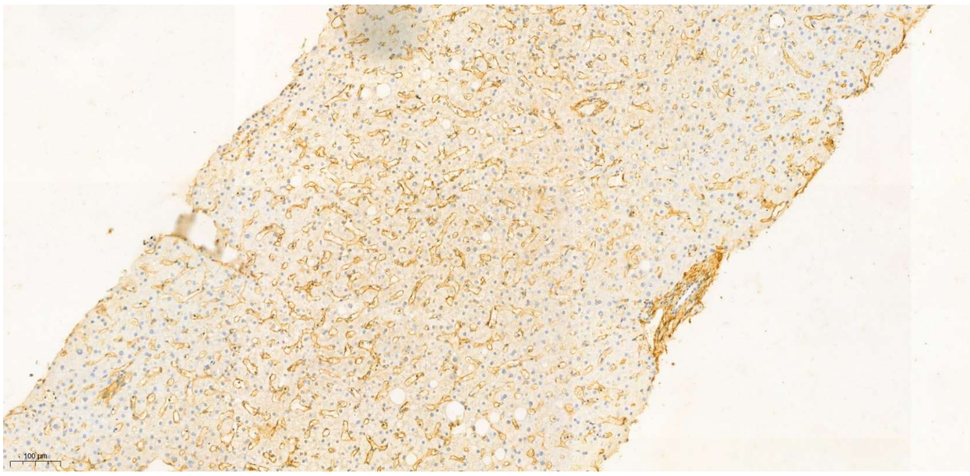


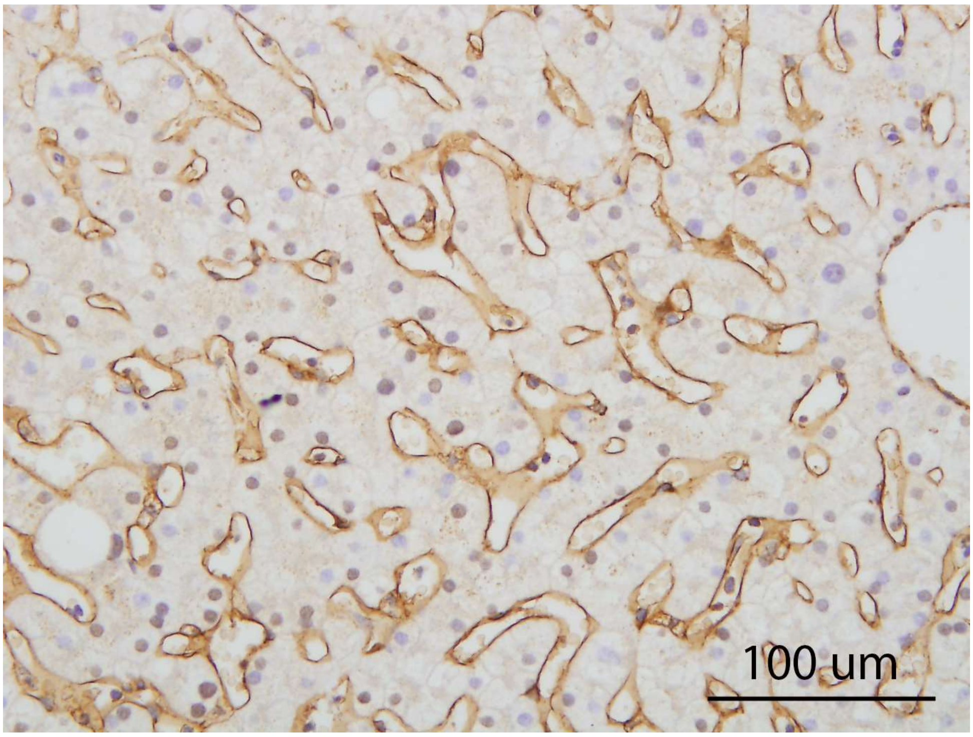
